# Engagement of health workers and peer educators from the National Adolescent Health Programme-Rashtriya Kishor Swasthya Karyakram during the COVID-19 pandemic: findings from a situational analysis

**DOI:** 10.1101/2022.03.28.22273059

**Authors:** Monika Arora, Stefanie Dringus, Deepika Bahl, Zoya Rizvi, Heeya Maity, Smritima Lama, Amanda J. Mason-Jones, Deepak Kumar, Prairna Koul, Shalini Bassi

## Abstract

**Background:** To understand the impact of COVID-19 on implementation of the peer education programme of the National Adolescent Health Programme-Rashtriya Kishor Swasthya Karyakram; repurposing of the RKSK health workers and Peer Educators (PE) in COVID-19 response activities and; adolescents′ health and development issues.

**Methods:** Virtual in-depth interviews were conducted with stakeholders (n=31) (aged 15 to 54) engaged in the implementation of the peer education programme at state, district, block, and village levels in Madhya Pradesh and Maharashtra (India). These interviews were thematically coded and analysed to address the research objectives.

**Results:** Despite most peer education programme activities being stopped, delayed, or disrupted during the pandemic and subsequent lockdown, some communication networks previously established, helped facilitate public health communication regarding COVID-19 and RKSK, between health workers, peer educators, and adolescents. There was significant repurposing of RKSK health workers and Peer Educators’ role towards COVID-19 response-related activities. Peer educators, with support from health workers, were involved in disseminating COVID-19 information, maintaining migrant and quarantine records, conducting household surveys for finding COVID-19 active cases and providing essential items (grocery, sanitary napkins, etc.) to communities and adolescents.

**Conclusion:** Peer Educators with support from community health workers are able to play a crucial role in meeting the needs of the communities during a pandemic. There is a need to further engage, involve and build the skills of Peer Educators to support the health system. PEs can be encouraged by granting more visibility and incorporating their role more formally by paying them within the public health system in India.

## Introduction

The COVID-19 pandemic has presented numerous challenges to health systems globally(1). India has implemented control measures similar to other countries to curtail the spread of COVID-19. This led to decision-makers making difficult choices, which involved prioritising preventive and curative COVID-19-related care while scaling back other areas of healthcare delivery, including but not limited to, adolescent health(2). The central and state government continue implementing preventive public health measures (3,4) including lockdowns, closure of educational institutions (schools, colleges), physical distancing, compulsory wearing of a mask, and restrictions on large gatherings and social events (deaths and marriages).

However, the pandemic has also had severe direct and indirect consequences on the health outcomes of people and the health system functioning during India’s first lockdown (March-May, 2020). Health care services were disrupted for non-COVID patient care due to the closure of routine outpatient departments, immunization clinics, antenatal services(5), and hospitals being designated as COVID-19 treatment centers(6). Data from National Health Mission highlighted a reduction in availing medical treatment (including both inpatients and outpatients) and emergencies for both infectious and non-communicable diseases (NCDs)(7) during the lockdown.

Implementation of several national health programmes, including India’s National Adolescent Health Programme-Rashtriya Kishor Swasthya Karyakram (NAHP-RKSK), were impacted during COVID-19. RKSK was launched across India in 2014, aiming to reach all adolescents, including both males and females, rural and urban, married and unmarried, and those in and out of school. RKSK encompasses various interventions involving multiple implementation stakeholders (Table 1). Interventions specified in Table 1 operate at school, community, and health facilities’ levels. The peer education programme, one of the core interventions under RKSK, is expected to increase adolescents’ engagement with health services and improve their knowledge, attitudes, and life skills in six thematic areas-mental health, injuries, and violence, sexual and reproductive health, NCDs, substance misuse, and nutrition.

**Table 1:**
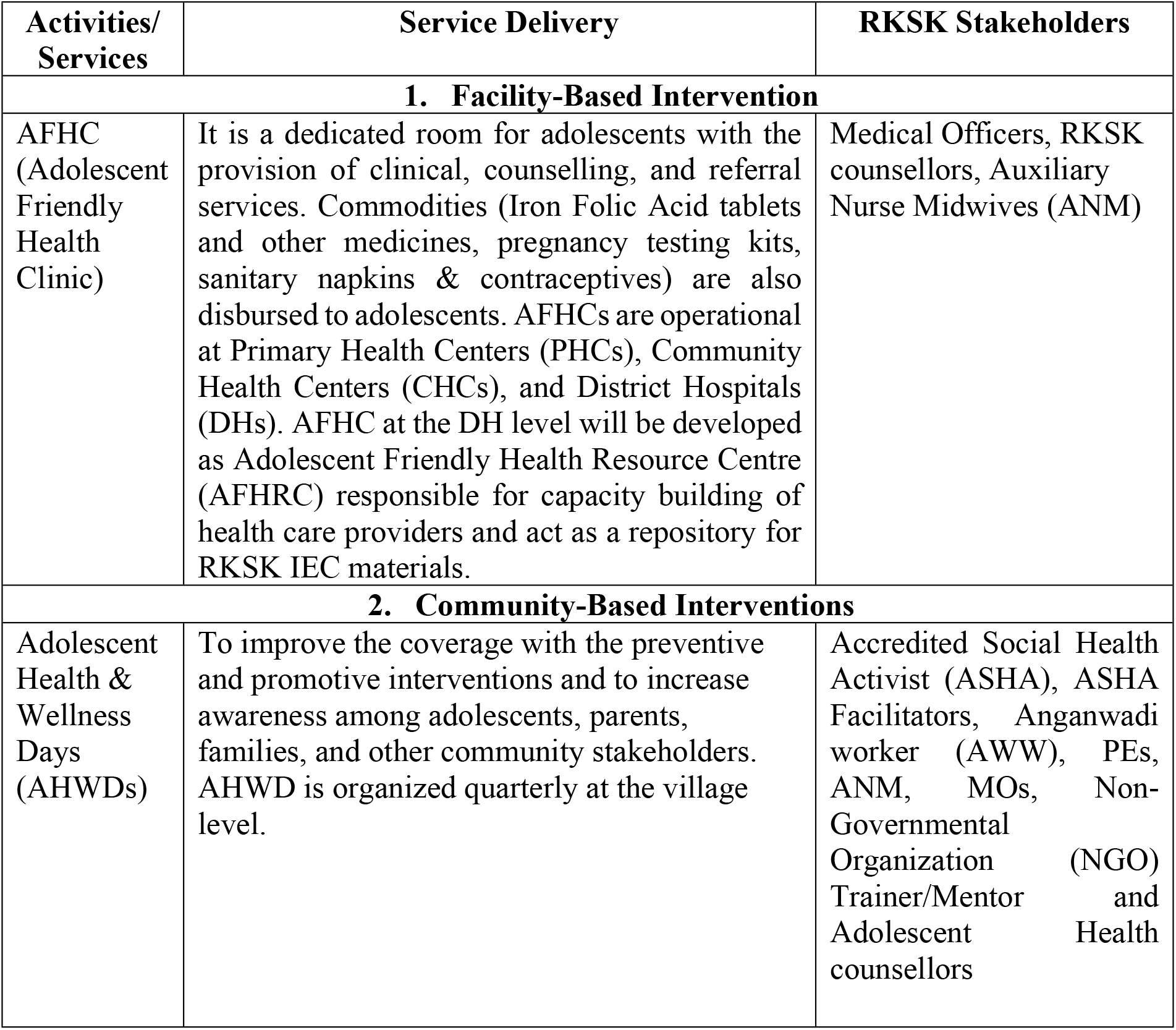
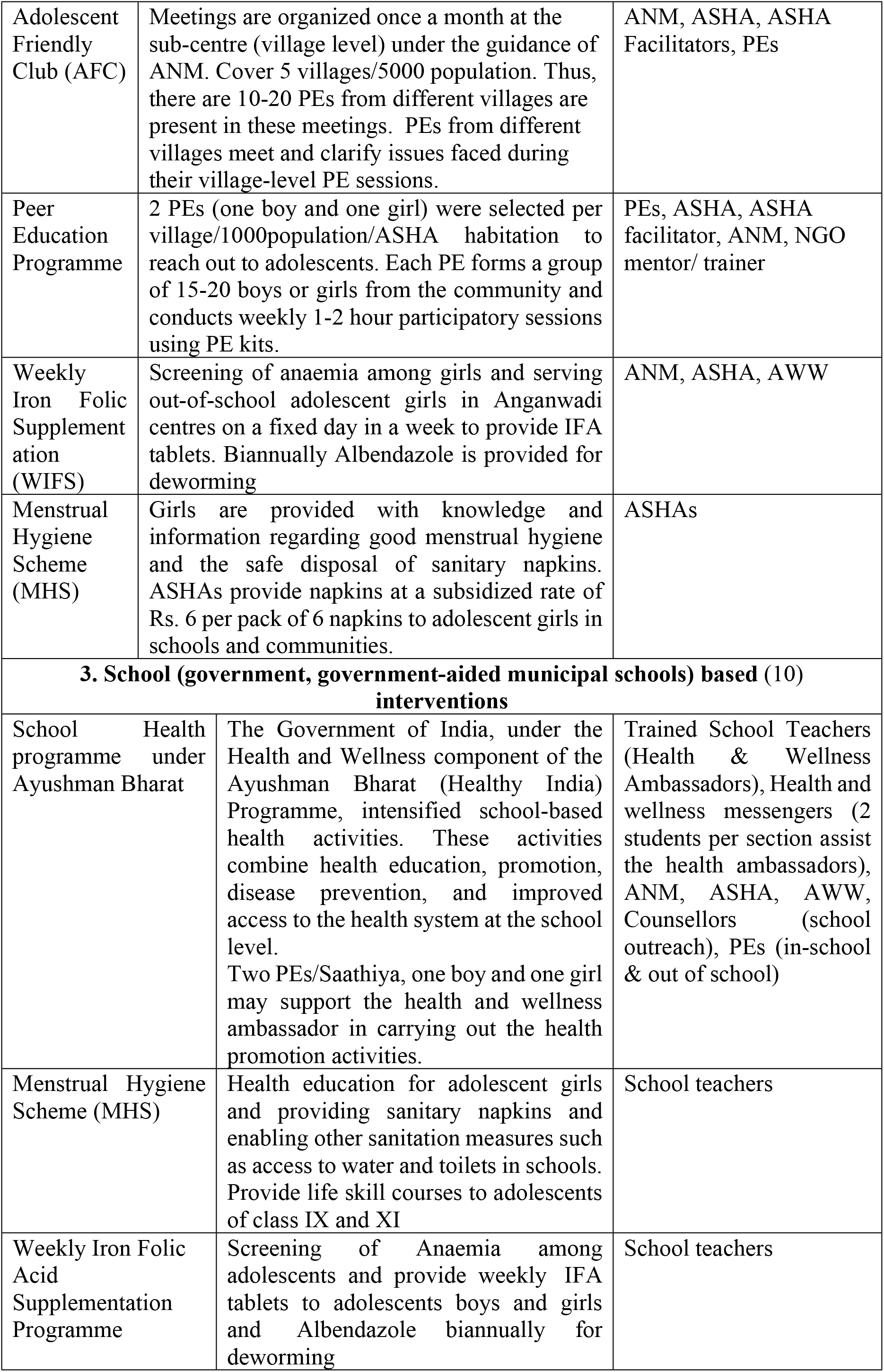
Overview of RKSK interventions and health workers (8,9) involved in programme implementation.

| Activities/<br>Services | Service Delivery | RKSK Stakeholders |
| --- | --- | --- |
| **1. Facility-Based Intervention** |  |  |
| AFHC<br>(Adolescent<br>Friendly<br>Health<br>Clinic) | It is a dedicated room for adolescents with the provision of clinical, counselling, and referral services. Commodities (Iron Folic Acid tablets and other medicines, pregnancy testing kits, sanitary napkins & contraceptives) are also disbursed to adolescents. AFHCs are operational at Primary Health Centers (PHCs), Community Health Centers (CHCs), and District Hospitals (DHs). AFHC at the DH level will be developed as Adolescent Friendly Health Resource Centre (AFHRC) responsible for capacity building of health care providers and act as a repository for RKSK IEC materials. | Medical Officers, RKSK counsellors, Auxiliary Nurse Midwives (ANM) |
| **2. Community-Based Interventions** |  |  |
| Adolescent Health & Wellness Days (AHWDs) | To improve the coverage with the preventive and promotive interventions and to increase awareness among adolescents, parents, families, and other community stakeholders. AHWD is organized quarterly at the village level. | Accredited Social Health Activist (ASHA), ASHA Facilitators, Anganwadi worker (AWW), PEs, ANM, MOs, Non-Governmental Organization (NGO) Trainer/Mentor and Adolescent Health counsellors |
| Adolescent Friendly Club (AFC) | Meetings are organized once a month at the sub-centre (village level) under the guidance of ANM. Cover 5 villages/5000 population. Thus, there are 10-20 PEs from different villages are present in these meetings. PEs from different villages meet and clarify issues faced during their village-level PE sessions. | ANM, ASHA, ASHA Facilitators, PEs |
| Peer Education Programme | 2 PEs (one boy and one girl) were selected per village/1000population/ASHA habitation to reach out to adolescents. Each PE forms a group of 15-20 boys or girls from the community and conducts weekly 1-2 hour participatory sessions using PE kits. | PEs, ASHA, ASHA facilitator, ANM, NGO mentor/ trainer |
| Weekly Iron Folic Supplementation (WIFS) | Screening of anaemia among girls and serving out-of-school adolescent girls in Anganwadi centres on a fixed day in a week to provide IFA tablets. Biannually Albendazole is provided for deworming | ANM, ASHA, AWW |
| Menstrual Hygiene Scheme (MHS) | Girls are provided with knowledge and information regarding good menstrual hygiene and the safe disposal of sanitary napkins. ASHAs provide napkins at a subsidized rate of Rs. 6 per pack of 6 napkins to adolescent girls in schools and communities. | ASHAs |
| **3. School (government, government-aided municipal schools) based (10) interventions** |  |  |
| School Health programme under Ayushman Bharat | The Government of India, under the Health and Wellness component of the Ayushman Bharat (Healthy India) Programme, intensified school-based health activities. These activities combine health education, promotion, disease prevention, and improved access to the health system at the school level.<br>Two PEs/Saathiya, one boy and one girl may support the health and wellness ambassador in carrying out the health promotion activities. | Trained School Teachers (Health & Wellness Ambassadors), Health and wellness messengers (2 students per section assist the health ambassadors), ANM, ASHA, AWW, Counsellors (school outreach), PEs (in-school & out of school) |
| Menstrual Hygiene Scheme (MHS) | Health education for adolescent girls and providing sanitary napkins and enabling other sanitation measures such as access to water and toilets in schools. Provide life skill courses to adolescents of class IX and XI | School teachers |
| Weekly Iron Folic Acid Supplementation Programme | Screening of Anaemia among adolescents and provide weekly IFA tablets to adolescents boys and girls and Albendazole biannually for deworming | School teachers |

Our research team initiated a study to explore the implementation of the peer education programme of RKSK in two Indian states (Madhya Pradesh and Maharashtra) in 2020. As part of this study, a situational analysis was conducted virtually, to examine the impact of COVID-19 on a) peer education programme implementation, b) repurposing of the RKSK health workers and PEs i.e. *in addition to their existing roles and responsibilities under the RKSK, their participation in COVID-19 response activities*, c) adolescents′ health and development issues.

## Materials and Methods

### Study setting and participants

The study was conducted in Madhya Pradesh and Maharashtra. Madhya Pradesh is a large state in central India with a population of 72.6 million and a literacy rate of 69.3%. Maharashtra, in India’s western peninsular region, is the second-most populous state with a population of 112.3 million and an 82.3% literacy rate(11). Both states were among the most severely affected states in India due to COVID-19(12,13). This trend remained from the start of the pandemic to its waning phase, with Maharashtra reporting an increasing number of cases despite the national lockdown(14). Madhya Pradesh, one month into the pandemic, remained among the top five most impacted states with the highest number of fatalities(15). In consultation with state health departments, two districts were chosen from each state, i.e., Panna and Damoh from Madhya Pradesh and in Maharashtra, Nashik, and Yavatmal, to include in the study. Study participants were recruited using a snowball sampling approach wherein the state RKSK team nominated participants from the district who then nominated participants from the block and further participants at the village level. Participants who provided their consent were included in the study.

### Data Collection

Data was collected through in-depth interviews (IDIs), using semi-structured interview guides tailored for each group of participants. These in-depth interview guides were developed referring to RKSK Implementation Guidelines(8), Operational Framework(16), and consultations with the Central, State, and District RKSK officials. An overview of respondents interviewed and themes covered has been presented in table S1. These IDIs were conducted in Madhya Pradesh (during June-November, 2020) and Maharashtra (during August-November 2020). IDIs were moderated by trained qualitative researchers accompanied by a note taker. To mitigate the risk of COVID-19 transmission, virtual audio interviews using the Zoom platform and mobile phones were conducted. Most officials at the district level were interviewed on Zoom while those at the village level were interviewed over the mobile phone considering the technology empowerment. The average duration of each interview was 60-90 minutes. Verbal consent was obtained from each participant as per the Indian Council of Medical Research’s revised guidelines during the COVID-19 pandemic (17). Interviews were conducted in English or Hindi, audio-recorded, translated, and transcribed. Ethics approval for the research was obtained from the Institutional Ethics Committee of the Public Health Foundation of India (Reference # TRC-IEC-342.1/17).

### Data Analysis

Each transcript was read several times by two authors independently, data were coded and organised thematically. Discrepancies between two authors in coding and organising data were discussed with a third senior author. In the end, three major and two sub-themes were generated to address the research objectives and these have been discussed in the results section.

## Results

### Demographic profile of study participants

In total, thirty-one participants were interviewed from both study states representing the RKSK at state, district, block, and village levels (Table 2). There were 18 participants from Madhya Pradesh and 13 from Maharashtra. The difference in the number of participants recruited by hierarchy (state, district, block, and village) and by states has been attributed to the nominations provided by the contact person at a higher level (state and district). The participants’ ages ranged from 15 to 54 years and the affiliation period with RKSK ranged from 1 month to 5 years while the median affiliation period was 2 years (24 months).

**Table 2:** Study participants & their roles and responsibilities (N=31)

|  | **RKSK health workers** | **Roles & Responsibilities** | **State** | **No. of Participants (N)** | **Participant Gender (Male/Female)** |
| --- | --- | --- | --- | --- | --- |
| **S. No.** | **State Level** |  |  |  |  |
| 1. | State Nodal Officer | Act as focal point person for adolescent health programme in the State.<br>Responsible for implementation of RKSK at State/ District Level.<br>Advocate for the importance of adolescent health at State<br>Coordination with all stakeholders to ensure appropriate implementation of the programme.<br>Prepare adolescent Health Programme Implementation | Maharashtra | 1 | M |
|  |  | Plan (PIP) for the State<br>• Publicity of adolescent health issues at different platforms using IEC, mass media, mid media, etc.<br>• Consolidate, review and submit the Adolescent Health Progress report every quarter to the Mission Director and further on to the Government of India<br>• Engage in regular monitoring and supportive supervision of progress of Adolescent Health Programmes. |  |  |  |
|  | **District Level** |  |  |  |  |
| 2. | Programme Manager /Chief Medical Officer (CMO) | • District CMO appoints officer to supervise, monitor, and report the implementation of RKSK<br>• Supervises, monitors, compiles, and reports on the activities of RKSK and NGO wherever applicable. | Madhya Pradesh | 1 | M |
| 3. | Community Mobilizer | • Supervises and monitors the ASHAs and ASHA Facilitators of the district for all<br>programs including RKSK components | Madhya Pradesh | 1 | M |
| 4. | Programme Coordinator | State government appointee and oversees RKSK implementation | Madhya Pradesh | 1 | M |
| 5. | District Reproductive and Child Health (RCH) Officer | Master Trainer of MOs, ANMs, ASHAs, PEs, and Lady Health Visitors<br>Monthly review of counsellor for monitoring the PE district improvement<br>Monitoring and evaluation of the programme | Maharashtra | 1 | M |
| 6. | District RKSK Counsellors | Dual responsibility of providing clinical and counselling services at Maitri clinic and District Hospital<br>Coordinates community and school-based outreach activities<br>Identifies and addresses the needs of adolescents to ensure support<br>Record Keeping & Reporting | Maharashtra | 2 | F |
| 7. | Counsellor at District Hospital | Provides clinical and counselling services<br>Coordinates community and school-based outreach activities | Madhya Pradesh | 1 | M |
| 8. | Training Faculty<br>(Faculty Training Centre) | • Monitor implementation of annual training plan monthly<br>• Conduct Training Visit according to the training plan and provide qualitative input.<br>• Coordinates and plans activities at the training centre<br>• Develop training plans and conducts training of trainers (MOs & other paramedical staff) according to the training plan<br>• Evaluation and feedback of the training conducted and gives input where necessary for improvement. | Maharashtra | 1 | F |
| 9. | NGO Representative | • NGO responsible for the recruitment of staff to support village and AFC level activities<br>• Report on activities and progress to District Nodal Officer | Madhya Pradesh | 1 | M |
| **Block Level** |  |  |  |  |  |
| 10. | Medical Officer<br>(Primary Health Centre) | • Dual responsibility of Medical Officer at PHC and counsellor at AFHC in PHC as RKSK coordinator for the block | Maharashtra | 1 | F |
|  |  | • Monitor AHD & AFC activities |  |  |  |
| 11. | RKSK Counsellor | • Counsellor at AFHC in CHC<br>• Provides counselling to adolescents on six themes of RKSK<br>• Supports district RKSK nodal officer to organize RKSK activities in their catchment area | Madhya Pradesh | 1 | F |
|  | **Village Level** |  |  |  |  |
| 12. | ANMs | • Moderates monthly Adolescent Friendly club meetings/sessions<br>• Providing counselling services at PHC<br>• Helps disburse non-financial incentives to PEs<br>• Organizes AHWD and provides group/individual sessions with parents on adolescent health needs<br>• Submits monthly progress report to PHC medical officer in charge | Maharashtra | 2 | F |
| 13. | ASHA Facilitators | • Supervises up to 10-12 ASHAs<br>• Attends at least one PE session per month to resolve issues raised by ASHAs/PEs<br>• Collects and consolidates PE<br>monthly reports<br>(collected by<br>ASHAs) | Madhya Pradesh | 2 | F |
| 14. | ASHAs | Helps select & recruit PEs in consultation with community leaders, parents, and NGO trainer-cum-mentor (where applicable)<br>Supportive supervision of PEs (help from groups, unresolved queries from PE sessions, etc.)<br>First point of contact for resolving issues faced by PEs<br>Attends PE sessions organized at village level<br>Collects and consolidates PE monthly reports<br>Helps organize, mobilize adolescents and other stakeholders to attend AHWD<br>Refers adolescents to AFHCs | Maharashtra<br>Madhya Pradesh | 2<br>5 | F<br>F |
| 15. | NGO Mentor cum Trainer | In states with NGO-led implementation models, they are appointed by NGOs<br>Helps in selection and recruitment of PEs along with ASHAs | Madhya Pradesh | 1 | M |
|  |  | Provides supportive supervision to PEs<br>Trains and distributes kits to PEs<br>Facilitates AFC meetings |  |  |  |
| 16. | Anganwadi Worker | Maintains Anganwadi Centre sometimes used for RKSK activities<br>Mainly engaged in WIFS component of RKSK services<br>Helps in organizing AHWD and mobilizing community participation | Madhya Pradesh | 1 | F |
| 17. | PEs | Conducts sessions with adolescents to build their knowledge, attitude, and skills on the six RKSK themes<br>Forms adolescent groups (brigade)<br>Provides referrals for AFHCs and/or counselling<br>Participate in the Adolescent Friendly Club meetings<br>Actively participate in Adolescent Health & wellness Days (AH&WD) | Maharashtra | 1 | M |
|  |  |  |  | 2 | F |
|  |  |  | Madhya Pradesh | 2 | M |
|  |  |  |  | 1 | F |
|  |  | • Maintains PE diary in the prescribed format<br>• Prepares monthly composite PE sessions' report |  |  |  |

Key findings from this situational analysis are described under the following themes and sub-themes: 1) implementation of Peer Education programme during COVID-19; a) connectivity between RKSK health workers and PEs and between PEs and adolescents; b) impact of COVID-19 on Adolescent Friendly Health Services (AFHCs) and helplines, and;2) repurposing of the RKSK health workers and PEs; 3) adolescents’ health and development issues during COVID-19.

### Implementation of Peer Education programme

In the face of the pandemic, all activities related to the Peer Education programme implementation were either on hold, significantly delayed, or altered. Disruptions in the programme implementation were due to the restrictions put in place due to COVID-19 (lockdown, physical distancing, restricted movements, etc.) The stalled activities ranged from signing contracts with Non-Governmental Organisations (NGOs) to implement programme activities such as Adolescent Friendly Club (AFC) meetings, recruitment and training of PEs, delivery of PE sessions, and community outreach activities such as Adolescent Health and Wellness Days (AHWDs), school outreach activities. The District Coordinator from Madhya Pradesh said “*due to COVID-19, the entire programme has been on pause, otherwise we would have received the copy of the agreement (between the state government and NGO) by now and started the work including staff selection. Due to a rise in the number of COVID-19 cases, we have PE training on hold”*. The District Reproductive and Child Health (RCH) officer in Maharashtra explained, “*we conduct outreach programmes in 5-6 schools every month but this year due to COVID-19, we haven’t been able to do it*”.

### Connectivity between RKSK health workers and PEs, and between PEs and adolescents during COVID-19 lockdown

Participants shared that PE sessions and information dissemination via in-person methods at the village level were put on hold due to COVID-19 restrictions. However, some communication, through mobile phones (via calls, WhatsApp, videos/ films) with PEs, related to COVID-19 and peer education programmes existed during the lockdown. Pre-existing WhatsApp groups for RKSK related activities, between Accredited Social Health Activists (ASHAs) and PEs, were used for this communication. This informal and unplanned communication was evidence of the pivot, from the original guidelines (18) and shift from PE programme implementation pre-COVID to accommodate needs emerging due to COVID context. This information was further shared with friends, family, and other adolescents.

> *We are giving information to PEs through WhatsApp only. All information given is related to coronavirus, like what precautions they should take, hand washing, wearing a mask when going out, cleaning used masks, maintaining a distance of at least 6 feet*. ***(ASHA, Madhya Pradesh*)**

In-person interaction with adolescents continued to occur during home visits by ASHAs and ASHA facilitators as part of the Weekly Iron Folic Acid Supplementation (WIFS) programme or other maternal and child health programmes.

Additionally, an NGO representative described how a trainer-cum-mentor, responsible for providing supportive supervision to PEs, had kept in touch with some PEs in Madhya Pradesh. He said, “*one mentor is in touch with 20 PEs through telephone. Since they (PEs) stay in the same village, they may talk to the adolescents one to one physically, or in small groups under the supervision of mentor over video call”*.

PE also reported sharing information about COVID-19 to other adolescents in their communities as instructed by ASHA:

> *We meet girls in the coaching classes and we talk about wearing a mask and hand washing. ASHA asked us to inform others about it*. ***(PE Female, Madhya Pradesh***)

Despite these successes of communication, some hindrances in the flow of information remained; for example, not all PEs could be reached via mobile phones due to lack of access. In many instances, there was sporadic and brief communication with those that were able to connect:

> *There are some pockets in our district where there is no network. Some adolescents don’t have their cell phones, some use their family member’s phone, so they may not be available to talk for that long when mentor calls*. ***(NGO Representative, Madhya Pradesh)***

### Impact of COVID-19 on Adolescent Health Services (AFHCs and Helpline)

In both states, participants reported adverse impacts on the delivery of health services, including AFHCs. During the initial lockdown period, AFHCs at Community Health Centres in Madhya Pradesh remained closed with no tele-counselling provision. At this time, counsellors were involved in other COVID-19 related duties. As reported by an AFHC counsellor at a CHC in Madhya Pradesh “*…the clinic is closed nowadays. We have to check migrant labourers; whether they have been screened or not for COVID-19. We collect data and send it to the concerned authorities. Nothing like that (tele-counselling etc*.*) is provided*.” The participants were also asked if there was any alternative available for adolescents such as a telephone helpline. There was a mixed response from the two study states participants. In Maharashtra, most of the participants were unaware of a helpline. A counsellor from Madhya Pradesh mentioned 1098, a 24-hour emergency outreach service helpline for children (0-18 years) in distress, a service of the Ministry of Women and Child Development operating through Childline India Foundation(20,21) and an active 104-helpline before COVID-19.

> “*Calling 104 was usually an effective way for adolescents to solve their problem when they may not feel comfortable speaking to an ASHA, as stay in the same village*.*” (****ASHA, Madhya Pradesh***)

However, the counsellor from CHC expressed concern about the inability to divert calls from 104, if adolescents require further or more individualised advice or medical care.

After the reopening of AFHCs in both states, the footfall of adolescents was reported to have decreased significantly. This decrease may have been attributed, in part, to parents’ hesitation in sending their children to clinics due to fear of COVID-19 transmission. In Maharashtra, a decline in footfall may also have reduced clinics’ functional days or vice-versa.

> *“Before COVID the clinic was regular (5 days a week). These days the footfall is much lesser, only 6-7 adolescents come in a day compared to 20 adolescents in a day before the pandemic*.*” (****Counsellor, District Hospital, Maharashtra)***

Due to COVID-19 restrictions, there may also have been gender differences in those who were accessing the services in clinics. A counsellor from AFHC reported (Maharashtra), “*before COVID-19 there would be 30-32 adolescents in a day and there were more female visitors than males. But there is not much difference now during COVID*.”

### Repurposing of RKSK health workers and PEs to support COVID-19 response: During and post-lockdown

Several respondents reported significant repurposing in terms of roles and responsibilities to support COVID-19 response-related activities during and post COVID-19 lockdown. The PEs who have been trained under RKSK adapted to the context and undertook COVID-19 response activities by disseminating COVID-19 information to community members including adolescents. With the intent of tracing and tracking, they helped maintain migration and quarantine records of people migrating from urban cities to villages, and records were submitted at the block level.

In Maharashtra, RKSK counsellors and counsellors of other health programs like Integrated Counselling and Testing Centers (ICTC) under the National AIDS Control Programme reported providing COVID-19 related counselling at help desks set up at out-patient departments and the in-patient department wards of District Hospitals. Similarly, from Madhya Pradesh, a counsellor from Community Health Centre (CHC) reported his involvement in checking the migrant labourers for COVID-19 symptoms.

> *“We have to check migrant labourers; whether they have been screened or not for COVID-19”. (****CHC Counsellor, Madhya Pradesh***)

ASHAs in Maharashtra were tasked with conducting village-level household surveys for the “My Village, My Responsibility” (19) campaign initiated by the Government of Maharashtra. The campaign aims to survey and screen households to detect COVID-19 patients and those with other pre-existing conditions (diabetes, cancer, and hypertension). A few ASHAs from Maharashtra also reported making and distributing masks using their resources (money, fabric, sewing machine) for community members at no charge. ASHA Facilitators and ASHAs were also tasked with visiting and surveying households to monitor COVID-19 active cases.

Without any additional formal training, PEs also contributed during COVID-19. They helped ASHAs in sensitising villagers about COVID-19 information including physical distancing, compulsory wearing of masks, sanitization, and handwashing. It was reported by the NGO trainer-cum-mentor (Madhya Pradesh) “*(PEs) are distributing masks, preparing migrant workers’ lists who are coming back and maintaining information about the people who are quarantined. PEs also helped ASHAs paint walls with information regarding COVID-19 preventive and protective measures. This included information about social distancing, masks, and sanitization and handwashing*.*”*.

In Maharashtra, PEs reported helping ASHAs in putting stickers as part of the “My Village, My Responsibility” (19) campaign.

> *As part of the My Village, My Responsibility campaign, we helped ASHA to put stickers outside the houses in the whole village. In that sticker, we write the number of family members, with their age, and if anyone has any comorbidities like diabetes, and if any member has a fever*. ***(PE Male, Maharashtra)***

PEs also provided essential items such as grocery and menstrual hygiene products to adolescents and/or their families in the villages living in containment (*only essential activities allowed with strict perimeter control ensuring no movement of people*) or red zones (*areas with high COVID-19 caseload*).

> *Those who were in red zones and not able to go out and buy anything. So, they call us (PEs) for necessities like milk, bread, and even medicines and we buy them and leave them outside their home*. ***(Male PE, Maharashtra)***

### Adolescents′ health and development issues during COVID-19

There was limited and mixed data on the potential impact of COVID-19 on adolescents’ health and development issues. Participants from both states shared that adolescents faced challenges pertaining to education due to COVID-19 related school closures.

> *“Their online classes are happening, and a lot of children are saying we don’t understand much in these online classes. I think in face-to-face teaching; they understand quickly in comparison to online classes. In our village, online study content is received on WhatsApp. According to adolescents, the information is so long that they lose interest in reading the text*.*”* ***(ASHA, Madhya Pradesh)***

Few interview participants reported that the adolescents did not face severe challenges like mental health or sexual abuse during the pandemic. PE also highlighted that mental health was the most discussed topic during PE sessions with adolescents at the village level before COVID-19.

> *Adolescents are wearing masks and meeting their friends, so I don’t think that COVID-19 is affecting their mental health*. ***(ASHA, Madhya Pradesh)***
>
> *The same number of cases of sexual abuse had come, both before and after COVID-19, only 2-4 cases came each month*. ***(Counsellor-District Hospital, Maharashtra)*** *Mental health is the most discussed topic among adolescents as I think they are more stressed, so they like sharing that with us*. **(PE, Maharashtra)**

PEs also reported that COVID-19 appeared to have had some impact on menstrual health hygiene. Two female PEs (Maharashtra) explained, *“sanitary pads/ napkins were not easily available. We contacted ASHA and ASHAs provided pads to the girls who were having difficulties in procuring them*.” There appeared to be a lack of demand in some places even pre-COVID, as reported by NGO trainer/mentor (MP) “*they very rarely use pads in the villages. Our NGO used to sell pads which cost rupees 15(USD 0*.*20). Few bought but most said that they feel better using cloth*.*”*

## Discussion

COVID-19, an unprecedented global health emergency, resulted in the disruption of healthcare delivery to people of all age groups, including adolescents. Literature suggests several consequences of COVID-19 and one of the most impacted groups would be the adolescents due to school closures, increased unemployment, etc. putting them at an increased risk of dropping out from school, gender gaps in education, mental health conditions, early marriage, smartphone addiction, etc. (1,22). To the best of our knowledge, this is the first study in India that has attempted to understand what happened to the national adolescent health programme (NAHP-RKSK) at the time of COVID-19 and how PE without any additional formal training, along with the health workers, played a crucial role in the COVID-19 response-related activities in the community of selected districts of Madhya Pradesh and Maharashtra.

### Implementation of Peer Education Programme

Findings from our study indicate that RKSK and its core interventions were put on hold or altered as efforts were diverted to COVID-19 response activities. Other countries(23) have also reported the diversion of their health systems to solely focus on COVID-19. RKSK and its core interventions aim to address adolescent health issues and if such interventions are hampered, it can affect development gains made in the past few decades(24) and negate improvements gained from previous maternal, child health, and nutrition-related national programs(25). For instance, disruption due to COVID-19 on programmes to end child marriage, coupled with the pandemic-caused economic downturn, could result in 13 million more child marriages over the next decade (2020-2030) across the globe (26).

Evidence from previous pandemics in sub-Saharan Africa and recent evidence from Ghana, Nigeria, and Kenya has revealed possible disparaging mid-and long-term impacts of COVID-19 on adolescents(23). Hence, in India, RKSK has a significant role to play in ensuring that developmental gains made in the area of adolescent health (in the field of nutrition (anaemia), SRH (teenage marriage, menstrual hygiene, use of contraceptives), violence (gender-based violence), etc.) are not reduced or lost.

#### Connectivity between RKSK health workers and PEs, and between PEs and adolescents during COVID-19 lockdown

In some instances, despite lockdown, RKSK health workers continued to provide informal support and supervision to PEs and adolescents by communicating about RKSK and COVID-19 precautions through phone calls, WhatsApp, or home visits. But there existed a digital divide in connecting with all PEs due to lack of mobile phones and/ or weak mobile networks. Some PEs maintained connections and support with adolescents in the community, either one-to-one or through video calls. At times group calls with mentors and PEs were also organised in Madhya Pradesh. These findings indicate that pre-established communication networks played a critical role in keeping adolescents, PEs, and RKSK workers connected, but also highlight the existence of a digital divide in most LMICs which has been brought to the forefront due to the pandemic(27). Further, there was evidence of the vital link that PEs play between community and health workers as iterated in the programme guidelines (18).

### Impact of COVID-19 on Adolescent Health Services (AFHCs & Helpline)

Our study data suggests access to health services was affected by the pandemic resulting in the closure of AFHCs, or reduction in clinic operating days, and diversion of the health workers to other COVID-19 related duties. A study reports that ASHAs’ and ANMs’ involvement in COVID-19 related response activities hampered the regular benefits ensured by RKSK such as the implementation of WIFS providing supplementary nutrition and provision of sanitary napkins(22). However, our findings indicate that ASHAs and ASHA facilitators as part of WIFS and maternal and child health programmes were making house visits and were talking to adolescents regarding the PE programme. It may be said that despite the pandemic, some efforts continued to reach out to adolescents. Moreover, we need these efforts to be sustained with clear job aids so that every adolescent in a village of India irrespective of location is benefitted uniformly.

As per our study, Madhya Pradesh already had a 104 toll-free helpline number that was widely used by adolescents before COVID-19. The Central Government of India introduced a free helpline (08046110007) to address psycho-social concerns due to COVID-19 (28) but the same was not reported from either of the two study states, despite data collection begun a few months after the introduction of the helpline. Literature encourages greater accessibility to teleconsultations and online psychotherapy with national helplines catering to mental health needs (29). These will be helpful during both the ongoing and future pandemics, helping adolescents to remain connected with services due to physical inaccessibility. Thus, COVID-19 has brought forward previously unrecognized healthcare service needs such as teleconsultations and toll-free helplines, (30).

### Repurposing of RKSK health workers and PEs to support COVID-19 response

There was a repurposing of the RKSK health workers and PEs towards COVID-19 response activities such as disseminating COVID-19 information, some instances of providing sanitary napkins, delivering essential groceries to the community, making and distributing masks, maintaining migrant and quarantine records, etc. These tasks were carried out despite challenges such as increased COVID-19 caseloads in both states, the absence of additional PE training, and no additional financial support to PEs. These are indicative of the fact that, despite the unpreparedness, the RKSK health workers and PEs were able to actively respond to the pandemic situation and provided help in whichever manner possible.

### Adolescents’ health and development issues during COVID-19

Our study states, the RKSK implementers interviewed perceived that there was no change/negative impact on the adolescents’ mental health. This could be attributed to various factors like the informal communication of health workers with PEs and adolescents, and mental health being the most commonly discussed theme during the PE sessions pre-COVID. Hence, adolescents may have been equipped with coping skills to manage the stress and anxiety due to the pandemic. Our situational analysis began during the initial phases of the COVID-19 pandemic and a truer picture would arise later due to the prolonged nature of the pandemic. However, the literature suggests COVID-19 could have a deleterious impact on the mental health of children and adolescents (31) and this may be due to a variety of reasons, varying from a low understanding of the entire situation, disruption in their regular schedule with school closures(32), problems of substance misuse, and heightened physical abuse (domestic violence, intimate partner violence)(33). These topics fall under the six RKSK themes and there is plenty of information available about the same in different resources such as the PE training manual, PE Activity Book, and Reference Book of Frequently Asked Questions (34).

India is already dealing with poor mental health care infrastructure and the added stress of the pandemic has overwhelmed the health care workforce with increasing pressure on the public health infrastructure(25). Studies discussed above indicate the need to focus on mental health as an upcoming health issue for both adolescent health programmes and in the larger context of public health. Therefore, PE interventions can be game-changers for the future of our country as they not only increase awareness but also encourage adolescents’ engagement as being beneficial for the overall health and well-being of adolescents (35).

Keeping in mind the growing need for mental healthcare in India(29) it may be suggested that the PEs as health navigators may help in providing support to the health system as a low-cost intervention as part of RKSK. Studies are indicating that in comparison to traditional health care providers, peer educators are more cost-effective and may help overcome shortages of human resources since community health workers are also overburdened(36,37).

Future program planning can be informed by our study findings to adopt a mix of high tech (digital platforms and tools), low tech (SMS and phone calls), as well as no-tech (community, teachers, and parents’ groups) approaches to reach more adolescents including PEs in a variety of contexts(27) while also assisting in overcoming the impact of future pandemics(38). Looking at the current context in India, PE sessions can inculcate the importance of physical distancing, general hygiene, and other infection control measures vital in preventing the transmission of viruses. Our study suggests that peer educators can be important stakeholders during a health disaster-related response, or a quick onset crisis, and in overworked resource-constrained health systems, especially in low-and middle-income countries like India. Hence, to keep the momentum and engagement of the PEs, there is a need to enhance the skills of Pes through booster trainings, ongoing support, granting them more visibility, and incorporating their new roles and responsibilities more formally within the health system(22). The PEs could also be provided experience certificates or some remuneration in the form of stipends for their work to acknowledge their contribution and for PEs to get a sense of accomplishment. This also empowers them at the community level and makes them responsible citizens within the health system(39).

Despite the novel findings presented in this paper, a few study limitations are worth considering. All interviews were audio-recorded thus participants’ non-verbal cues could not be captured, which could have enriched descriptions of experiences and situations in the study. Due to snowball sampling the number of participants at state, district, and village levels were not equal. Thus, there could be reporting bias by the participants due to their difference in the level of engagement with programme implementation. In context to the impact of COVID-19 on adolescent health and development issues, we have captured the perspective of various stakeholders other than PEs. As indicated by the stakeholder, there was a limited impact so we did not explore these issues further by gender, socio-economic status, marital status, etc. We could not include adolescents who are part of RKSK in this study, thus the beneficiary perspective is missing. Finally, our study was limited to a few participants from two states, limiting our findings’ generalizability. All the above limitations will be considered while conducting an ongoing larger study with a bigger set of participants in the same study sites.

## Conclusion

Our findings indicate that there were some avenues of access to health care available to the adolescents despite the pandemic. They also highlight how RKSK health workers and PEs who were already working in the community were repurposed to engage with the COVID-19 response to not only the adolescents but also their communities as a whole. PEs along with the health workers navigated this challenging time and demonstrated their resilience in the face of an emergency. While their roles may have been very critical in responding to the pandemic, some additional support in the form of trainings, compensation, etc. could help strengthen this model for the future.

## Data Availability

The data that support the findings of this study are available on request from the corresponding author. The data are not publicly available due to privacy or ethical restrictions. As per our Institutional data sharing policy, prior approval is required from the Research Management Committee (RMC) and Principal Investigator (PI) of the study. After the Committee?s approval on the request, de-identified data can be shared only for publication.

## Acknowledgments

We would like to express our sincere gratitude to all the respondents of our study. We also acknowledge the help and support of various officials from the district, state health departments, and the Ministry of Health and Family Welfare, Government of India. We also thank Dr. David Ross for critically reviewing the manuscript being a member of the Independent Project Steering Committee formulated for the study. The corresponding author also affirms that she has listed everyone who has contributed significantly to this study.

## Supporting Information

S1 Table: An overview of respondents interviewed and themes covered

## References

1. OECD. Youth-and-COVID-19-Response-Recovery-and-Resilience -OECD. 2020.

2. World Health Organization & United Nations Children’s Fund (UNICEF). Community-based health care, including outreach and campaigns, in the context of the COVID-19 pandemic: interim guidance, May 2020. 2020 May.

3. Guidelines on the measures to be taken by Ministries/ Departments of Government of India, State/ Union Territory Governments and State/ Union Territory Authorities for containment of the COVID-19 Epidemic in the Country [Internet]. 2020 [cited 2020 Dec 3]. Available from: https://www.mha.gov.in/sites/default/files/Guidelines_0.pdf

4. MHA Order with Addendum to Guidelines Dated 24.03.2020 [Internet]. [cited 2020 Dec 3]. Available from: https://www.mha.gov.in/sites/default/files/MHAorderwithaddendumtoGuidelinesDated24.3.2020_0.pdf

5. Garg S, Bhatnagar N, Singh Mm, Borle A, Raina S, Kumar R, et al. Strengthening public healthcare systems in India; Learning lessons in COVID-19 pandemic. J Fam Med Prim Care. 2020;9(12):5853.

6. Dutta SS. 600 more hospitals in India designated as Covid-19 treatment centres within four weeks-The New Indian Express. Indian Express. 2020;

7. Arora M. COVID-19 and Beyond: Implications for People Living with Non-Communicable Diseases in India | Center for the Advanced Study of India [Internet]. CENTER FOR THE ADVANCED STUDY OF INDIA. 2020 [cited 2021 Feb 15]. Available from: https://casi.sas.upenn.edu/iit/monikaarora

8. Ministry of Health and Family Welfare. Implementation guidelines Rashtriya Kishor Sawasthya Karyakram(RKSK). GoI. 2018.

9. National Health Mission (NHM) G. Operational framework -translating strategy into programmes. 2014BC.

10. Development R, Welfare F. Ministry of Health and Family Welfare Government of India Operational Guidelines on School Health Programme under Ayushman Bharat Health and Wellness Ambassadors partnering to build a stronger future A Joint initiative of Ministry of Health “; Family W. 2018;(April).

11. Kumar S, Kumar S. Use of cluster analysis to monitor novel coronavirus-19 infections in Maharashtra, India. Indian J Med Sci. 2020 Aug;72(2):44–8.

12. Rath RS, Dixit AM, Koparkar AR, Kharya P, Joshi HS. COVID-19 pandemic in India: A Comparison of pandemic pattern in Selected States. Nepal J Epidemiol. 2020 Jun;10(2):856–64.

13. Ghosh P, Ghosh R, Chakraborty B. COVID-19 in India: Statewise Analysis and Prediction. JMIR Public Heal Surveill 2020;6(3)e20341 https://publichealth.jmir.org/2020/3/e20341. 2020 mAug;6(3):e20341.

14. KRISHNAN SANJANA, DEO SAHIL, MANURKAR SHARDUL. 50 days of lockdown: Measuring India’s success in arresting COVID-19 | ORF. 2020 May.

15. Coronavirus India lockdown Day 33 updates | April 26, 2020 -The Hindu.

16. Adolescent Health Division, Ministry of Health and Family Welfare G of I. Rashtriya Swasthya Kishore Karyakram Operational Framework. 2014.

17. Mathur R. National Guidelines for Ethics Committees Reviewing Biomedical & Health Research [Internet]. Indian Council of Medical Research. 2020 [cited 2021 Apr 29]. Available from: https://www.icmr.gov.in/pdf/covid/techdoc/EC_Guidance_COVID19_06052020.pdf

18. Adolescent Health Division M, GoI. Operational Framework, translating strategy into programmes.2014 Jan.

19. Press Trust of India. ‘My family, my responsibility’ drive will make Maharashtra fit: CM. Hindustan Times [Internet]. 2020 Sep 25 [cited 2021 Apr 29]; Available from: https://www.hindustantimes.com/india-news/my-family-my-responsibility-drive-will-make-maharashtra-fit-cm/story-OH3cJ1CjjT2n5MSmEv4dUM.html. Published September 25, 2020. Accessed January 31, 2021.

20. Poddar S, Mukherjee U. Ascending Child Sexual Abuse Statistics in India During COVID-19 Lockdown: A Darker Reality and Alarming Mental Health Concerns. Indian J Psychol Med. 2020 Sep;42(5):493–4.

21. Khurana Manju, Dayal Aditi. Childline 1098 (Case Studies on E-Governance in India 2013-14). Delhi;

22. Kumar Mm, Karpaga Priya P, Panigrahi S, Raj U, Pathak V. Impact of COVID-19 pandemic on adolescent health in India. J Fam Med Prim Care. 2020;9(11):5484.

23. Addae EA. COVID-19 pandemic and adolescent health and well-being in sub-Saharan Africa: Who cares? Int J Health Plann Manage. 2021 Jan;36(1):219–22.

24. Ghosh A, Nundy S, Mallick TK. How India is dealing with COVID-19 pandemic. Sensors Int. 2020 Jan;1:100021.

25. Gopalan HS, Misra A. COVID-19 pandemic and challenges for socio-economic issues, healthcare and National Health Programs in India. Diabetes Metab Syndr Clin Res Rev. 2020 Sep;14(5):757–9.

26. UNFPA. Impact of the COVID-19 Pandemic on Family Planning and Ending Gender-based Violence, Female Genital Mutilation and Child Marriage. Interim Technical Note. 2020.

27. C3. ISSUES FACED BY ADOLESCENTS DURING COVID19. 2020 Sep.

28. MoHFW. COVID-19 Helplines for Mental Health Assistance. 2021.

29. Banerjee D, Bhattacharya P. “Pandemonium of the pandemic”: Impact of COVID-19 in India, focus on mental health. Psychol Trauma Theory, Res Pract Policy. 2020 Sep;12(6):588.

30. Kumar A, Rajasekharan Nayar K, Koya SF. COVID-19: Challenges and its consequences for rural health care in India. Public Heal Pract. 2020 Nov;1:100009.

31. Dalal PK, Roy D, Choudhary P, Kar SK, Tripathi A. Emerging mental health issues during the COVID-19 pandemic: An Indian perspective. Indian J Psychiatry. 2020 Sep;62(Suppl 3):S354.

32. Imran N, Zeshan M, Pervaiz Z. Mental health considerations for children & adolescents in COVID-19 Pandemic: Pakistan J Med Sci. 2020 May;36(COVID19-S4):S67–72.

33. Population Foundation of India. Impact of COVID-19 on young people: Rapid assessment in three states, May 2020 (Bihar, Rajasthan and Uttar Pradesh) [Internet]. 2020. Available from: https://populationfoundation.in/wp-content/uploads/2020/08/Rapid-Assessment_Report_Youth_Survey_Covid.pdf

34. Peer Educator Reference Book-FAQs.

35. Efuribe C, Barre-Hemingway M, Vaghefi E, Suleiman AB. Coping With the COVID-19 Crisis: A Call for Youth Engagement and the Inclusion of Young People in Matters That Affect Their Lives. J Adolesc Heal. 2020 Jul;67(1):16–7.

36. Sikander S, Lazarus A, Bangash O, Fuhr DC, Weobong B, Krishna RN, et al. The effectiveness and cost-effectiveness of the peer-delivered Thinking Healthy Programme for perinatal depression in Pakistan and India: The SHARE study protocol for randomised controlled trials. Trials. 2015 Nov;16(1):1–14.

37. He J, Wang Y, Du Z, Liao J, He N, Hao Y. Peer education for HIV prevention among high-risk groups: A systematic review and meta-analysis. BMC Infect Dis. 2020 May;20(1):1–20.

38. MUKHERJEE DEBARATI, BEHAL SHREYA, KURIAN OOMMEN C. Investing in Adolescent Health: Harnessing India’s Demographic Dividend | ORF. ORF. 2020.

39. Bernays S, Tshuma M, Willis N, Mvududu K, Chikeya A, Mufuka J, et al. Scaling up peer-led community-based differentiated support for adolescents living with HIV: keeping the needs of youth peer supporters in mind to sustain success. J Int AIDS Soc. 2020 Sep;23(S5):e25570.

